# Epidemiology of the use of inferior vena cava filters in Brazil between 2008 and 2019

**DOI:** 10.1101/2021.08.23.21262490

**Authors:** Andressa Cristina Sposato Louzada, Dafne Braga Diamante Leiderman, Marcelo Fiorelli Alexandrino da Silva, Maria Fernanda Cassino Portugal, João Carlos de Campos Guerra, Marcelo Passos Teivelis, Nelson Wolosker

## Abstract

**Purpose:** to study the totality of inferior vena cava filters placed in the Brazilian Public Health System, which insures more than 160 million Brazilians, trends by geographic region and in-hospital deaths after filter placements and also due to pulmonary embolism between 2008 and 2019.

**Patients and methods:** public and open data regarding in-hospital mortality due to pulmonary embolism and all inferior vena cava filters placed in Brazilian public hospitals between January 2008 and December 2019 were extracted from the Brazilian Public Health System’s databases.

**Results:** 9108 inferior vena cava filters were placed, 98.18% of which was therapeutic. There was an overall increasing use of these devices, statistically significant for all Brazilian regions, except the North. In-hospital mortality rate in patients who received inferior vena cava filters was 6.21%, stable over time. We observed an upward trend in in-hospital mortality rate due to pulmonary embolism, statistically significant for all Brazilian regions, except the North.

**Conclusion:** We observed a low standardized rate of inferior vena cava filter placements in Brazil between 2008 and 2019, but with a trend towards an increasing trend use. Almost all indications for filter placement were therapeutic. In hospital mortality in patients receiving inferior vena cava filters was high, 6.21%, and stable over time, whereas the trend of in-hospital mortality rate due to pulmonary embolism is increasing. Our findings were heterogeneous across Brazilian regions and contrasted to those observed in the USA, which is likely due to cultural and socioeconomic factors.

## Introduction

Inferior vena cava (IVC) filters were designed to prevent recurrent pulmonary embolism (PE) and/or decrease PE-related mortality. They have been placed since the 1960s(1), and this procedure is considered safe(2).

Its indication in patients with venous thromboembolism (VTE) and contraindication to anticoagulation therapy is well established(3–5). However, its use as an adjunct to anticoagulation in patients with VTE and its prophylactic use, that is, in patients with risk factors for thrombosis but without VTE, has been a topic of debate in the literature(6). The main guidelines vary in their recommendations on IVC filters’ indications(6), fostering a significant variability in clinical practice, with temporal fluctuation and regional differences.

Concerning temporal fluctuation, some authors have assessed the use of IVC filters and their trends in United States of America’s (USA) using the National Inpatient Sample and Medicare databases and they observed a consistently increasing use of IVC filters up to 2008(4,7–10), which was especially faster after the Food and Drug Administration (FDA) approval of retrievable filters in 2003, raising concerns about overuse in the literature(8). Between 2008 and 2010, when CHEST Guideline recommended against the prophylactic use of IVC filters(11) and the FDA issued a safety alert, after 921 device-related adverse events were reported(7), the rates of IVC filter placement stopped growing. From 2010 on, the use of IVC filter began to steadily decline until 2012 to 2015, years in which these studies ended(4,7,10,12,13).

Regarding regional differences, some authors reported that the variability between centers, states and regions could not be fully explained only by the regional differences in patients’ characteristics or in the incidence of VTE, suggesting that cultural and socioeconomic factors may play an essential role in the IVC filter placement(10,14).

To the best of our knowledge, no study evaluated the nationwide trends of IVC filter placements after 2015, therefore the effects of the renovation of the FDA safety alert in 2014(15), and the last CHEST Guideline in 2016(16) that stressed out their positioning against IVC filter prophylactic use, if any, are still unknown. In addition, there is no nationwide study evaluating these temporal trends and regional differences in a developing country.

Therefore we designed this research to study the totality of filters placed in public hospitals under Brazilian Public Health System, which exclusively insures most of the population, approximately 160 million Brazilians, and the trends of IVC filter use by geographic region between 2008 and 2019. Secondary objectives were to evaluate in-hospital mortality rates due to pulmonary embolism and in-hospital mortality rates after IVC filter placement during the study period.

## Material and methods

This is a retrospective population-based study that analyzed publicly available data from the TabNet platform, of the Department of Informatics of the Unified Health System (DATASUS)(17) and Oswaldo Cruz Foundation (Fiocruz)(18). The TabNet system provides open and anonymous data regarding hospital admissions and procedures performed in hospitals duly accredited with the Brazilian public health system, necessary for hospitals to be reimbursed by the government.

Statistics referring to IVC filter placement were selected at the TabNet platform of the Oswaldo Cruz Foundation (Fiocruz). Among the selections, the analysis included the number of IVC filters placed, using the single code assigned to this procedure in the platform of the management system for procedures and medications of the Brazilian public health system (04.06.04.014-1), the underlying diagnosis justifying the procedure as coded by the International Classification of Diseases, Tenth Revision, (ICD-10) and corresponding in-hospital mortality, grouped by Brazilian geographic region and distributed over the years 2008 to 2019.

Statistics referring to in-hospital deaths due to pulmonary embolism (ICD-10 I26) were selected at the TabNet platform of DATASUS, section hospital mortality, with the data being grouped by Brazilian geographic region and distributed over the years 2008 to 2019.

All data were collected from public access sites through computer programs for automated content access (web scraping). These automated navigation codes were programmed in Python language (v. 2.7.13, Beaverton – Oregon – USA) using the Windows 10 Single Language operational system.

The data collection, platform field selection, and table adjustment steps were performed using the selenium-webdriver packages (v. 3.1.8, Selenium HQ, several collaborators worldwide) and pandas (v. 2.7.13, Lambda Foundry, Inc. e PyData Development Team, New York, USA). The Mozilla Firefox browser (v. 59.0.2, Mountain – California – USA) and geckodriver webdriver (v 0.18.0, Mozilla Corporation, Bournemouth, England) were used. Following collection and treatment, all data were organized and grouped in a spreadsheet using Microsoft Office Excel 2016® (v. 16.0.4456.1003, Redmond – Washington – USA) software.

Statistical analysis: Linear regression was performed to evaluate the trends in the distribution of IVC filter placement and deaths due to pulmonary embolism among Brazilian regions throughout the years, using the SPSS® (IBM Corp. Released 2013. IBM SPSS v 22.0, Armonk, NY – USA). The established level of statistical significance was less than 0.05.

The Ethics Committee of the Hospital Israelita Albert Einstein approved it (CAAE: 35826320.2.0000.0071). Since we used a publicly available database with de-identified entries, a waiver of informed consent forms was requested and granted by the Institutional Review Board.

## Results

A total of 9,108 IVC filters were placed in the Brazilian Public Health System between 2008 and 2019. Considering that approximately 160 million inhabitants depend exclusively on the Brazilian Public Health System, this is equivalent to an average of 47.6 procedures per 10 million populations per year. As for the indications for ICV filter placements, 98.18% of the procedures were associated with an ICD-10 code of venous thromboembolism.

The distribution of IVC filter placement rates by Brazilian regions per 10 million inhabitants exclusively dependent on the public health system per year is shown in Figure 1. Overall, in Brazil, there was a statistically significant upward trend in IVC filters placement between 2008 and 2019 (p<0.001), which was also observed for all Brazilian regions, except the North, the region with the lowest rates of IVC filter use. The most expressive growth was observed in the South region, where the standardized rate of use of IVC filter in the last year of the study was twice the initial standardized rate.

**Figure 1.**
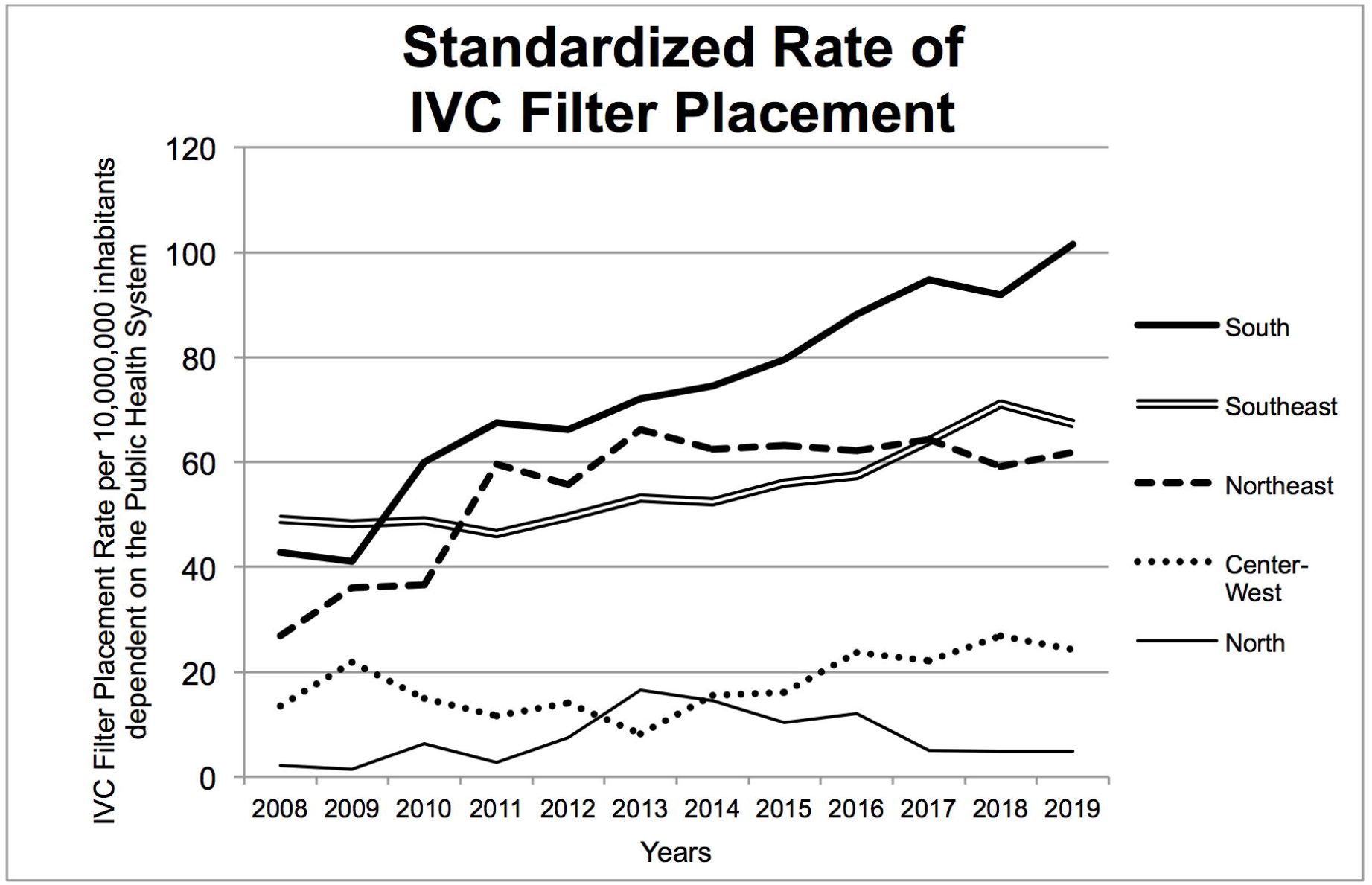
Distribution of IVC filter placement rates by Brazilian regions per 10 million inhabitants exclusively dependent on the public health system per year.

Distribution of in-hospital deaths following the placement of IVC filter is depicted in Figure 2. In total, there were 566 in-hospital deaths registered in patients receiving IVC filter in the study-period, with overall in-hospital mortality rate was 6.21%. We observed no statistically significant trend towards an increase or decrease in in-hospital mortality after IVC filter placement in any Brazilian region or overall in Brazil.

**Figure 2.**
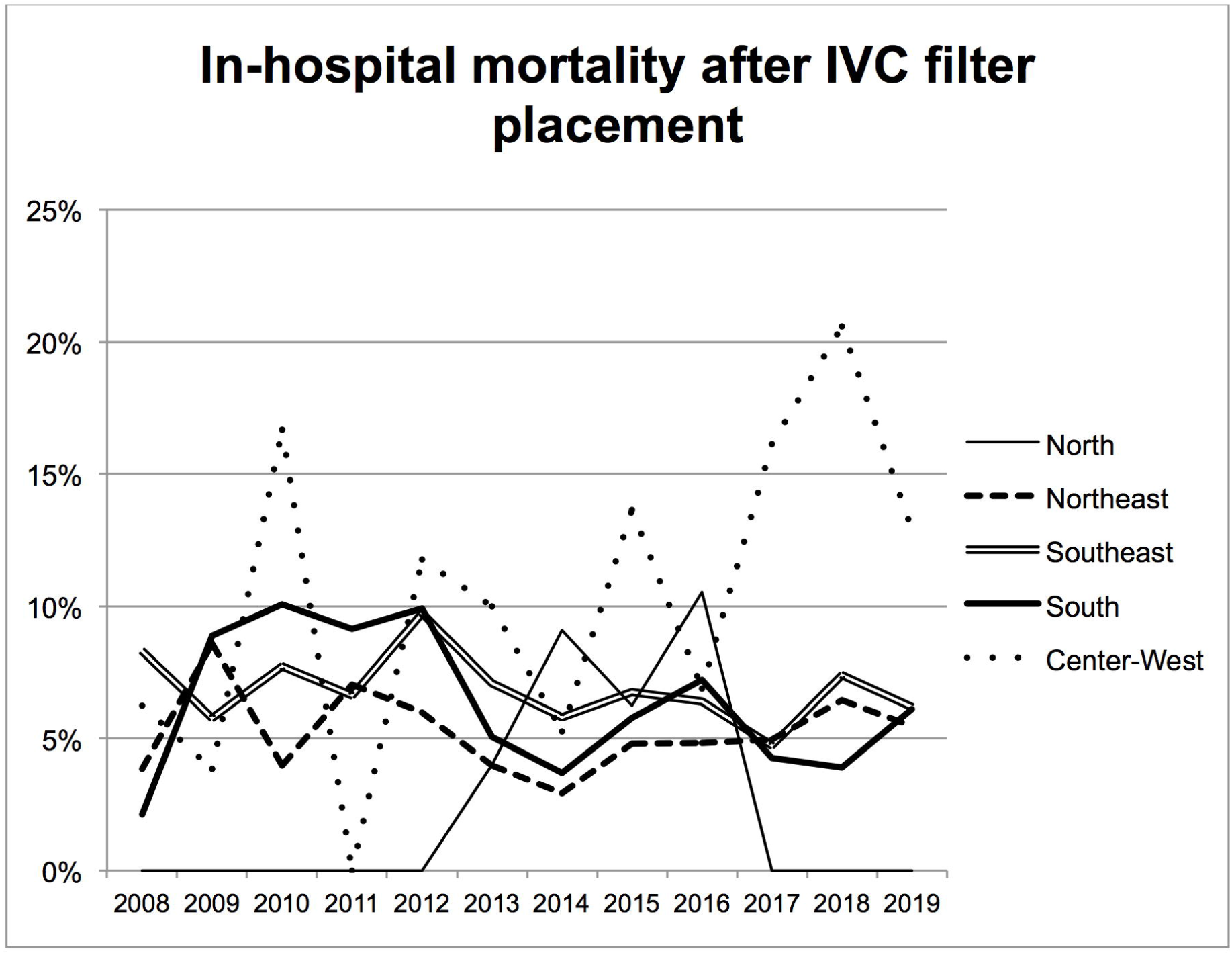
Distribution of in-hospital deaths following the placement of IVC filter among Brazilian regions between 2008 and 2019.

The distribution of total standardized rates of in-hospital mortality due to pulmonary embolism in public hospitals between 2008 and 2019 is shown in Figure 3. Overall, in Brazil, there was a statistically significant upward trend in in-hospital mortality due to pulmonary embolism from 2008 to 2019 (p<0.001), which was also observed for all Brazilian regions, except the North again.

**Figure 3.**
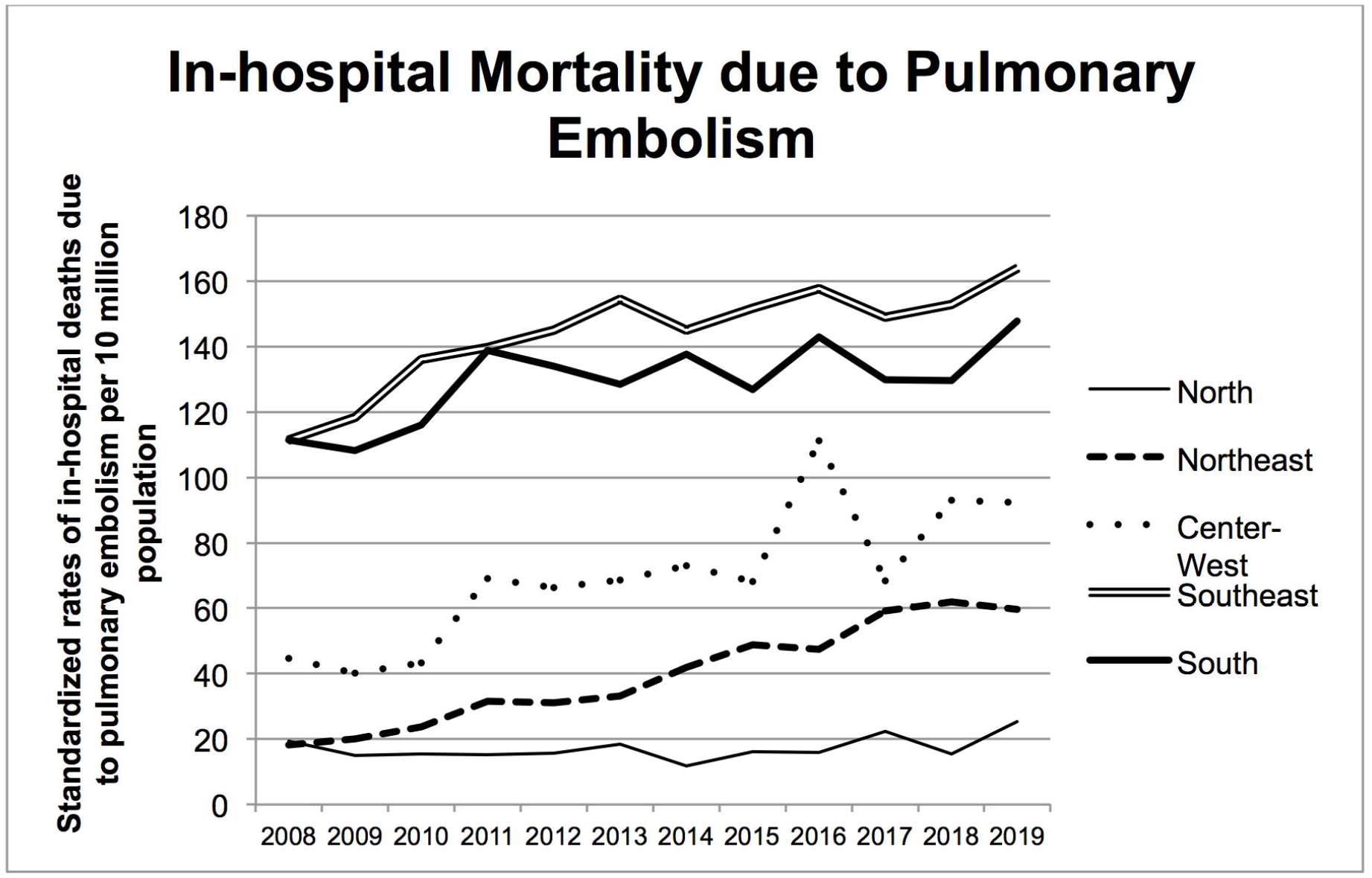
Distribution of total standardized rates of in-hospital mortality due to pulmonary embolism in public hospitals by Brazilian regions between 2008 and 2019.

## Discussion

### IVC filter placement rates

We observed a total of 9,108 IVC filter placements in Brazilian public hospitals, with an overall average of 47.6 procedures per 10 million populations per year, a rate that is up to 100 times lower than that in the USA(19) and 6 times lower than what was estimated for France, Germany, Italy, Spain and the United Kingdom, when analyzed in combination, in the late 2000s(6). Even when comparing rates after the decrease in the trends of IVC filter placement in the USA, the National Inpatient Sample registered 96,005 IVC filter placements in 2014, which is more than 10 times higher than the total observed in Brazilian public hospitals during twelve years(10).

Added to this disparity between the use of IVC filter in Brazil, a developing country with a relatively young publicly funded health system facing underfunding(20), and in developed countries as the USA and the European countries mentioned above, we observed an internal inequality, among Brazilian regions. For instance, in 2019, the rate of IVC filter placement in the South, a richer and more developed region(21), was 20 times higher than in the North, the region with the lowest rates of IVC filter placements, which is also the region with the worst development indexes(21), facing the greatest shortage of health professionals and facilities(22), and with the lowest population density(21) and difficult access to health care. Both these external and internal discrepancies underline that cultural and socioeconomic factors may have impact on the placement of IVC filters, which is in line with other reports(10,14).

### Trends in IVC filter use

The combination between a relatively low use of IVC filters and the observation that 98.18% of the IVC filters was placed in patients with VTE may explain the upward trend in IVC filter use over the years without any negative effects due to the FDA alerts or the CHEST guidelines. Since almost every indication of IVC filter was therapeutic, as opposed to a proportion of approximately 80% of the indications observed in the USA(7), recommendations against the prophylactic use of IVC filters should not impact the use of filter in Brazilian public hospitals, in contrast to what was observed in the USA(4).

### Mortality data

Along with an increasing trend of IVC filter use, we observed a stable in-hospital mortality rate after IVC filter placement and an increasing standardized rate of in-hospital mortality from pulmonary embolism.

The overall in-hospital mortality rate in patients receiving IVC filters was as high as 6.21%, which is similar to other reports in the literature(5,23). Rather than a procedural complication, this in-hospital mortality rate reflects the clinical severity of presentation of the patients who received the IVC filters. Unfortunately, we do not have access to patients’ demographics and comorbidities, which are known to strongly affect mortality in these cases, such as age, presence of pulmonary embolism associated with acute respiratory syndrome or shock(24), advanced stage cancer(25,26) and severe trauma(27), which is a limitation of our study. Nonetheless, the stable distribution of deaths following IVC filter placement over the years, observed in all regions, indicate that even though more IVC filters are being placed recently, the profile of the patients who are receiving them remains similar in terms of severity.

Data regarding the increasing overall in-hospital mortality due to PE underlines the relevance of this disease and the need to learn about the epidemiology of its treatments, which includes the epidemiology of the use of IVC filters. Similar to other studies, we failed to observe a positive impact on PE-related mortality associated with the increased use of IVC filter(28–30).

### Limitations

As with any database-driven study, the accuracy of the findings is susceptible to inherent coding or data entry errors.

Since data are anonymous, follow up was not possible. Therefore, we do not know the patients’ demographics, comorbidities and severity of presentation, as well as the causes of death. We were also unable to assess hospital incidence of PE, whether the patients who received IVC filters also received anticoagulation, type of IVC filter inserted and retrieval rates. In addition, we were unable to evaluate if trends and mortality were affected by hospital characteristics, as observed by other authors^15,32^.

Despite all limitations, our study provides comprehensive population-level findings in a developing country with a Public Health System that exclusively insures more than 160 million Brazilians. To the best of our knowledge, there is no nationwide study evaluating these temporal trends and regional differences in a developing country, which is essential, as we observed that data on the use of IVC filter in developed countries may not be generalized to all countries.

If every country could carry out this type of analysis, studying their total numbers through the Big Data assessment, in order to better plan its public health care policies, medical education and the allocation of resources, this could provide a more standardized care regarding the use of IVC filters and other types of medical devices.

## Conclusion

We observed a low standardized rate of inferior vena cava filter placements in Brazil between 2008 and 2019, but with a trend towards an increasing trend use. Almost all indications for filter placement were therapeutic. In hospital mortality in patients receiving inferior vena cava filters was high, 6.21%, and stable over time, whereas the trend of in-hospital mortality rate due to pulmonary embolism is increasing. Our findings were heterogeneous across Brazilian regions and contrasted to those observed in the USA, which is likely due to cultural and socioeconomic factors.

## Data Availability

This is a retrospective population-based study that analyzed publicly available data from the TabNet platform, of the Department of Informatics of the Unified Health System (DATASUS) and Oswaldo Cruz Foundation (Fiocruz)

## Disclosure

The author reports no conflicts of interest in this work.

## Notes

### Competing Interest Statement

The authors have declared no competing interest.

### Funding Statement

No funding was received.

